# Epidemiology and Associated Risk Factors of Diabetic Retinopathy in Patients with Diabetes Mellitus Attending a Tertiary-care Hospital in Hargeisa, Somaliland

**DOI:** 10.64898/2026.03.29.26349614

**Authors:** Hassan A. Yousuf, Ayesha S. Abdullah, Mohammad Shamsal Islam, Mulusew Asferaw Melesse, Asamere Tsegaw Woredekal

## Abstract

**Purpose:** Diabetic retinopathy (DR) is one of the most important complications of diabetes mellitus (DM), representing the leading cause of blindness among working age adults in developed countries. This study was aimed to investigate the epidemiology and risk factors of DR in patients with diabetes mellitus in a hospital setting in Somalia.

**Methods:** The study was an observational, descriptive cross-sectional and hospital-based study and data were collected from January 2023 to May 2023. A structured questionnaire was used to collect relevant demographic and clinical data. Both univariate and bivariate tables were used for analysis. Data analysis included frequency distribution, cross-tabulation, co-relation and association, and statistically significant tests between variables (X2, p-value, and CI).

**Results:** A total of 384 DM patients were studied and 76% (n=293) of them had type 2 DM. The average duration of diabetes mellitus was 9.7± 6.9 years and the mean age was 47.24 ± 19.36 years (range 18 -100 years old). A majority 66% (n=253) were female, about a third of them had normal body mass index (BMI) (n=172, 44.8%) and 170 (44.3%) had concomitant hypertension. About 51% of the patients (n=197) had DR out of which 17% had non-proliferative diabetic retinopathy (NPDR) (n=67) and 26% had Macular oedema (n=98). Age above 40 years (p=0.020), marital status (P=0.010), employment status (P=0.002) and literacy status (P=0.020) were significantly associated with the presence of DR. Patients aged below 40 had 37% lesser risk of having diabetic retinopathy than patients aged above 40 years. Longer duration of diabetes (p=0.001) and the presence of concomitant cardiac illness (p=0.001) were strongly associated with the presence of diabetic retinopathy. Patients with duration of diabetes more than 10 years had approximately 2 times higher chance of developing DR than those with duration less than 10 years.

**Conclusion:** The very high prevalence of DR (51%) among our patients implies the needs for a good health policy to manage DM and DR patients in Somalia. Effective regular eye screening and treatment for all diabetes patients should get priority.

## Introduction

Diabetic retinopathy is a leading cause of blindness in people of working age in developed countries and also emerging as one of the causes of blindness in developing countries. (1) Although effective treatments are available, their success depends on early detection and timely referral. (2) Diabetic retinopathy screening effectively reduces the risk of vision loss; however, screening attendance is consistently below recommended levels. (3)

Diabetes Mellitus is considered as a global epidemic because currently around 463 million (2019) people are suffering from DM and it’s expected that the figure will reach to 700 million by 2045. (4,5) Out of 1.5 billion people live with blindness and .4 million of them become blind from Diabetic Retinopathy (DR) globally. (6) Worldwide visual impairment has decreased, however, blindness induced by DR increased from 0.2 million to 0.4 million and moderate-severe visual impairment from 1.4 million to 2.6 million from 1990 to 2015. (7)

The global annual incidence of DR varies from 2.2% to 12.7% and the risk factors for the development of DR include long duration of diabetes, poor glycemic control, hypertension, hyperlipidemia, genetic predisposition and overweight. (8,9,10)

Some recent studies suggested that DR is increasingly becoming a major cause of blindness in Asia-Pacific and Africa continent. The prevalence of DR varies from 7% to 62% in African countries and there are no genuine large-scale statistics on the prevalence of diabetic retinopathy (DR) in Somalia and Somaliland. (11) However, a small community-based study reveals that DR prevalence in Mogadishu is 21.1%. (12)

This study was aimed to investigate the epidemiology and risk factors of DR in patients with DM who visited a tertiary eye hospital in Hargiessa, Somaliland.

## Materials and Methods

A descriptive cross-sectional study was conducted in a hospital setting. A structured and pre-tested questionnaire was used to collect data from the DM patients attending a tertiary hospital. A total of 384 DM patients were included in this study following the Fisher et al. 1991 formula. Study data were collected from January 2023 to May 2023. All consecutive Type 1 and Type 2 DM patients of age 18 and above who visited the Manhal Multispecialty Hospital during the study period were included.

### Clinical Examination and Data Collection

An experienced optometrist measured Presenting and Best Corrected Visual Acuity (BCVA) and Intraocular level (IOP). Patients’ pupil was dilated using tropicamide 1% eye drop. Anterior and posterior segment examinations were done using slit-Lamp and 90D Volk lens by senior ophthalmologists. Both Optical Coherence Tomography (OCT) and Fundus Examination were conducted to determine the stages of DR and the level of damage of macular edema due to DR was graded by senior ophthalmologists. Staging of the retinal changes was made using the Diabetic Retinopathy Study guidelines and recorded and subcategorized as Non-proliferative diabetic retinopathy (mild, moderate, and severe), Proliferative diabetic retinopathy (early, high risk, and advanced) and Diabetic macular edema (center involved, non-centered involved). (13) The average of Patients’ last three months Fasting Blood sugar (FBS) level was collected and for those with no record of the last three months FBS, HbA1C test was done and data registered. Weight and height were recorded, Body Mass Index (BMI) calculated and registered.

### Quality Control and Data Management

The study data were collected over 5 months period (January 2023 to May 2023) using an interview schedule that included the following variables: socioeconomic, demographic, risk factors of DM and DR, and Clinical Information. The study format and interview schedule were given to ophthalmologists for determining content validity. Their comments were incorporated in finalizing the research instruments. The filled-in formats and interview schedules were edited immediately after the collection of data every day. Again, data were cleaned before analysis.

## Data Analysis

Collected data was first edited and coded and then entered into the computer for analysis. Both univariate and bivariate tables were used for analysis. The data were analyzed using the Statistical Package for Social Science (SPSS). Data analysis included frequency distribution, cross-tabulation, co-relation and association, and statistically significant tests between variables (X2, p-value, and CI). Associations between DR and continuous and categorical variables were computed using Fisher’s exact test and Pearson χ2 test. Odds ratios (ORs) and 95% confidence intervals (CI) were used to determine the strength of the association between independent and dependent variables. All independent variables were cross-tabulated with dichotomized outcomes of DR (yes/ no). Simple binary logistic regression followed by multiple binary logistic regression was analyzed and conducted to identify factors associated with DR in the study population.

## Ethical Consideration

Institutional Review Board (IRB) was obtained from the School of Post-Graduate Studies and Research and Manhal Specialty Hospital, University of Hargeisa, Somalia. Written and verbal consent was obtained from each participant after explaining the purpose and nature of the research. Participation in the study was voluntary and participants were informed of their right to quit/refuse their participation at any stage of the study if they do not want to participate. Moreover, the confidentiality of the information was assured by using an anonymous consent form.

## Results

The study revealed that out of 384 DM patients 51% (n=197) had DR out of which 17% had NPDR (n=67) and 26% had Macular oedema (n=98). Two-third or 66% (n=253) were females and 131 (36%) were males with female to male ratio of 2:1.

Around 76% of DM patients had type-II diabetes mellitus (n=293) and the remaining 91 (24%) had type-I DM. The average duration of diabetes mellitus was 9.7± 6.9 years.

About 40% of DM patients were Illiterate, 28% had primary education, 21% had secondary level education and 10% had undergraduate level education. About 57% of DM patients were married and the majority of respondents were housewives and did not have any formal occupation.

Over two-thirds (70%) of them were with in age range between 40 to 80 years old and the mean age was 47.24 ± 19.36 years (range 18-100). Sixty two percent (n=57) of type-I DM patients were within age group between 10-21 years and the most frequent age group for type-II DM patients was between 51-60 years which accounted 32% (n=96) of them. We observed that Type-II diabetes mellitus increases with the patient’s age (x2=25.20, df=9, Sig.=0.001).

Body Mass Index (BMI) was with in normal range in 44.8% (n=172) of study patients and170 (44.3%) had hypertension, 28 (7.3%) had renal disease and 36 (9.4%) had cardiac disease. About 67% of patients had visual acuity in the normal range in their worst eye.

Demographic and clinical factors such as age, education level, residence, occupation, type of diabetes mellitus, presence of hypertension, presence of renal disease, presence of cardiac disease, body mass index, marital status, duration of diabetes mellitus and mode of treatment were tested to see the variables’ statistical association with diabetic retinopathy. (Table-I and II)

**Table I.**
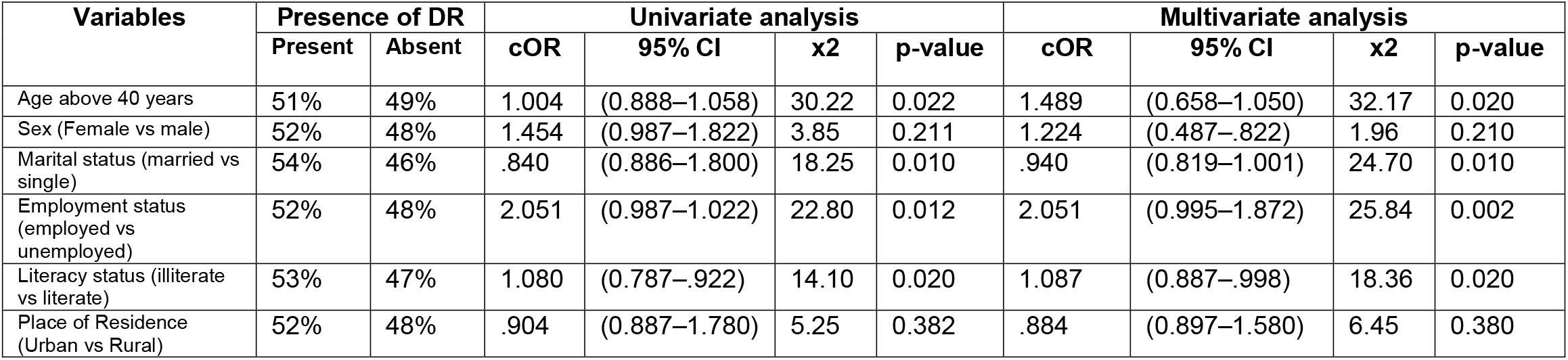
Socio-demographic factors and their association with the presence of DR among DM patients seen at Manhal Specialty Hospital, Hargiessa Somaliland, January 2023 to May 2023.

| Variables | Presence of DR |  | Univariate analysis |  |  |  | Multivariate analysis |  |  |  |
| --- | --- | --- | --- | --- | --- | --- | --- | --- | --- | --- |
|  | Present | Absent | cOR | 95% CI | x2 | p-value | cOR | 95% CI | x2 | p-value |
| Age above 40 years | 51% | 49% | 1.004 | (0.888–1.058) | 30.22 | 0.022 | 1.489 | (0.658–1.050) | 32.17 | 0.020 |
| Sex (Female vs male) | 52% | 48% | 1.454 | (0.987–1.822) | 3.85 | 0.211 | 1.224 | (0.487–.822) | 1.96 | 0.210 |
| Marital status (married vs single) | 54% | 46% | .840 | (0.886–1.800) | 18.25 | 0.010 | .940 | (0.819–1.001) | 24.70 | 0.010 |
| Employment status (employed vs unemployed) | 52% | 48% | 2.051 | (0.987–1.022) | 22.80 | 0.012 | 2.051 | (0.995–1.872) | 25.84 | 0.002 |
| Literacy status (illiterate vs literate) | 53% | 47% | 1.080 | (0.787–.922) | 14.10 | 0.020 | 1.087 | (0.887–.998) | 18.36 | 0.020 |
| Place of Residence (Urban vs Rural) | 52% | 48% | .904 | (0.887–1.780) | 5.25 | 0.382 | .884 | (0.897–1.580) | 6.45 | 0.380 |

**Table II.**
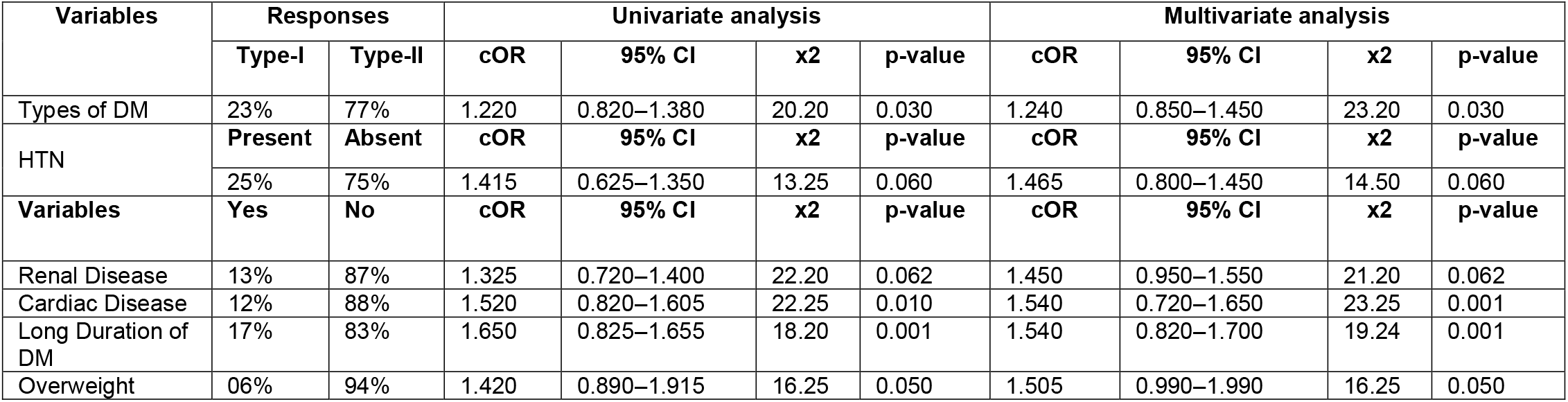
Clinical risk factors and their association with DR among DM patients seen at Manhal Specialty Hospital, Hargiessa Somaliland, January 2023 to May 2023.

The results of binary logistic regression analysis showed that only age (P=0.01) and the duration of diabetic mellitus (P=0.030), were found to be associated with the presence of diabetic retinopathy. Patients aged below 40 years had 37% less risk of the presence of diabetic retinopathy than patients aged above 40 years. (Table-I and II)

Among the demographic factors, age above 40 years (P=0.022), marital status (P=0.010), and literacy status (P=0.020) had significant association with the presence of diabetic retinopathy.

Longer duration of diabetes (p=0.001) and the presence of concomitant cardiac illness (p=0.001) were strongly associated with diabetic retinopathy. The presence of diabetic retinopathy with the duration of diabetes more than 10 years were approximately 2 times higher than duration of diabetes less than 10 years.

## Discussion

This is a cross-sectional study conducted at a tertiary-level hospital in Hargiessa, Somaliland, and it is the first report of its kind in this part of Somalia. The data of a total of 384 patients was collected and analyzed.

The study showed that the prevalence of diabetic retinopathy in these patients was 51.3% which is comparable to reports from Addis Ababa Ethiopia,51%, (14) and Zambia, 52%, (15) but higher than other reports from Ethiopia, 42%, (16), 34%, (17), Tanzania, 27.90%, (18), Egypt, 24.00%, (19) and Zimbabwe, 28.40%, (20).

The high prevalence seen in our study could be due to the fact that the sample population was taken from a referral eye hospital where most of the patients were referred from the medical diabetic clinic unlike some of the studies mentioned above which were done at medical diabetic clinics. Different sampling techniques, sample size, and diagnostic method may have also contributed to this difference. The disparities in the socioeconomic status of study patients in these countries and availability of health care facilities for diabetic care might have also affected the observed difference.

A majority of our study population were Type 2 DM patients (76%), a trend similar to studies done in Ethiopia, 88.4% (16), 53.6% (20), and 72.8% (19) and elsewhere in Africa where a majority of study patients had type 2 DM. (21,22,23,24,25)

The mean age of diabetes patients in this study was 47.2-/+ 19.3 years which is nearly comparable to a study done in Ethiopia, 45 years, (17) but lower than another Ethiopian study 55.4 years, (16) and studies in other parts of the world. (21,22,25,)

The female preponderance seen in our study patients, 66%, is in contrary to reports from Ethiopia where males accounted for 60% (16), 54.7% (17), 54% (26) of study patients. It has been said that men in developing countries tend to be economically more independent and able to visit health care facilities better than women (17). On the other hand, females may be more prone to diabetes mellitus as they are less involved in harder physical activities than men.

Our study showed longer duration of diabetes (p=0.001) was strongly associated with the occurrence of diabetic retinopathy. The presence of diabetic retinopathy with the duration of diabetes more than 10 years were approximately 2 times higher than duration of diabetes less than 10 years. This finding was consistent with major global meta-analyses and most of the studies done in other African countries. (16,17, 23,24,25,26,28)

We also found in this study that increasing age is associated with development of diabetic retinopathy (P=0.02) and our patients aged below 40 years old had 37% less risk of the presence of diabetic retinopathy than patients aged above 40 years. This trend is seen in many similar studies globally (,27,28,29,30) and an example is a study done in Ethiopia that reported the odds of developing diabetic retinopathy was six times higher in patients with age 60 years or older than in those younger than 60 years (30).

Age is a risk factor for diabetic retinopathy because the retina is more susceptible to damage over time due to blood vessels becoming more fragile and more likely to leak to cause diabetic retinopathy. (30)

Strangely, the presence of concomitant hypertension in our patients was not associated with the development of diabetic retinopathy although hypertension is an established risk factor for diabetic retinopathy and reported in many epidemiologic and laboratory-based studies. (10,24,31,32,)

The relatively modest sample size, the eye clinic based (hospital based) nature of the study which might have increased the prevalence of diabetic retinopathy and the absence of laboratory data to correlate with diabetic retinopathy are some of the limitations of this study.

## Conclusion

The high prevalence of diabetic retinopathy in this study implies the need for good health care policy to manage diabetes mellitus and diabetic retinopathy in this part of Somalia. Effective regular eye screenings for all diabetes mellitus patients for diabetic retinopathy should get priority.

## Data Availability

All data produced in the present study are available upon reasonable request to the authors

## Acknowledgment

We deeply thank our participant patients for their willingness to participate in the study and also Manahal Hospital for allowing us to conduct the study.

## Authors contributions

All authors together conceived and developed the study under the lead of the first author. All authors contributed for the development of the first draft and the entire team discussed the results and contributed to the final manuscript. All authors read and approved the final draft of the manuscript.

